# From non-uptake during early adolescence to acceptance during a catch-up campaign: a qualitative study of changing HPV vaccination decisions across two life stages among young women in Amsterdam, the Netherlands

**DOI:** 10.64898/2026.10.27.26366415

**Authors:** Sarah van Veelen, Puck Ellemijn Kahlmann, Laura Denis, Johannes Antonie Bogaards, Gertrude, Udi Davidovich, Eline Saskia Wijstma, Anouk Wisse, Maarten Schim van der Loeff, Catharina Johanna Alberts

## Abstract

**Background:** Human Papillomavirus (HPV) vaccination effectively prevents HPV-related disease, but its population-level impact depends on sufficient coverage. In 2023-2024, the Netherlands offered free HPV vaccination to individuals aged 19-27 years through a national catch-up campaign. For women, this was a second opportunity, as they had been offered vaccination at ages 9-13 through the National Immunisation Programme (NIP). Back then, uptake was relatively low (46-57% in these birth cohorts), but uptake increased to 55-63% following the catch-up vaccination. This campaign provided an apt opportunity to explore the factors underlying previous and current HPV vaccination decisions among the same women across two life stages.

**Methods:** Between May 2023 and May 2024, 16 semi-structured interviews were conducted in Amsterdam with women who accepted HPV vaccination during the catch-up after non-uptake during early adolescence. Interviews explored factors influencing non-uptake and later acceptance, guided by the Health Belief Model and additional sociocultural constructs. Data were analysed using thematic analysis with open and theory-guided coding in MAXQDA.

**Results:** Non-uptake during early adolescence was primarily shaped by parental decision-making, cultural and religious norms, fear of side effects, low perceived susceptibility to HPV and limited self-efficacy. Acceptance in young adulthood was driven by greater autonomy, higher perceived susceptibility, increased trust in accumulated vaccine evidence and accessible vaccination infrastructure. Peer influence replaced parental influence as the dominant social factor.

**Conclusion:** HPV vaccine decision-making is a dynamic process rather than fixed. Permanent catch-up HPV vaccination opportunities would enable young adults to make autonomous vaccination decisions as their circumstances change.

**Highlights:**

- Adolescent HPV non-uptake was driven by parents, culture and safety fears;
- Adult acceptance was driven by perceived risk, greater trust and peer influence;
- Decision-making is a dynamic state, not a fixed one-time decision;
- Free, accessible infrastructures to vaccination enabled catch-up uptake;
- Catch-up HPV vaccination enables autonomous vaccine decisions in young adults.

## Introduction

Human papillomavirus (HPV) is a highly prevalent sexually transmitted infection, with most sexually active individuals acquiring at least one HPV infection during their lifetime[1]. While the majority of infections clear spontaneously, persistent infection with high-risk HPV types can result in cervical, anogenital and oropharyngeal cancers[2]. HPV-related disease represents a substantial global public health burden that can be reduced with vaccination.

HPV vaccination is highly efficacious in preventing HPV infection and related disease, including strong evidence of protection against cervical cancer[3–6]. Vaccine effectiveness is highest when vaccination occurs before sexual onset, but vaccination in young adulthood can still provide protection depending on prior HPV exposure and future risk of infection[7]. Achieving population-level impact requires both high effectiveness and high uptake. As part of the cervical cancer elimination strategy, the World Health Organization aims for 90% HPV vaccination coverage among girls aged 15 by 2030[8]. However, HPV vaccination coverage remains well below this target globally[9]. Understanding opportunities to increase HPV vaccine uptake remains essential to improve coverage.

In the Netherlands, HPV vaccination was introduced in 2009 through the National Immunisation Programme (NIP), initially targeting girls aged 13 years. Since 2022, boys have also been included and the target age has been lowered to 9-10 years. Uptake was low among the first invited birth cohorts (1993–1996) in 2009, at 50%, but gradually increased to 65% in the most recently invited cohort in 2025[10]. Despite this improvement, uptake remains lower than that of other childhood vaccinations[10]. Between 2023 and 2024, the Netherlands implemented a one-year 18+ catch-up campaign offering free HPV vaccination to individuals aged 19–27 years (birth cohort 1996-2003). At initial invitation during early adolescence, uptake was relatively low in these birth cohorts (46-57%). After catch-up vaccination, uptake increased to 55-63% in these birth cohorts[11]. For women, this campaign provided a second opportunity to receive HPV vaccination, as they had already been invited during early adolescence. This context offered a unique opportunity to explore how vaccination decisions change over time within the same individuals.

Studies on HPV vaccination decision-making during early adolescence have identified high parental influence, low perceived risk of HPV due to the child’s young age, insufficient information and scepticism about the safety and necessity of HPV vaccines as key factors for vaccination non-uptake[12–15]. Determinants of vaccination decision-making in adulthood have been studied less extensively than those during early adolescence. Existing studies among young women who are offered vaccination at an older age identified low perceived risk of HPV infection, insufficient information and fear of side effects as reasons to refuse vaccination[16, 17]. However, little is known about individuals who initially did not get vaccinated during early adolescence but later accepted vaccination in adulthood. Understanding this transition may provide important insight into how vaccination decisions evolve across the life course.

In this qualitative study, we explore the factors that influenced young women’s decisions to accept HPV vaccination during the Dutch 18+ catch-up campaign, as well as their retrospective reflections on non-uptake during early adolescence. By specifically including young women who changed from adolescent non-uptake to vaccine acceptance in young adulthood, this study provides insight into factors associated with change in perceptions and vaccine decision-making over time. This study may help inform future vaccination strategies, communication efforts and policy decisions aimed at improving HPV vaccine uptake across the life course.

## Methods

### Setting and participants

From May 2023 through May 2024, Dutch men and women aged 19-27 years (birth cohort 1996 through 2003) who were unvaccinated or partially vaccinated against HPV were eligible for vaccination with the bivalent HPV vaccine (Cervarix) free of charge through Public Health Services within the Netherlands (18+ catch-up campaign). For men within this age range, this catch-up campaign marked their first opportunity to get vaccinated against HPV. For women, it was their second opportunity, as they had been offered HPV vaccination as a child during the Dutch NIP.

This study aimed to understand changes in decision-making processes and decisions at first NIP invitation during early adolescence compared with current decisions to vaccinate during the 18+ catch-up campaign. Therefore, this study focuses exclusively on women in birth cohorts 1996-2003, who had not taken up HPV vaccination via the initial NIP and who subsequently decided to receive HPV vaccination during the 18+ catch-up campaign. From May 2023 to May 2024, participants were recruited at the HPV vaccination sites in Amsterdam or via snowball sampling.

### Data collection

Sixteen semi-structured interviews were conducted using an interview guide, to obtain insights into the factors influencing the women’s vaccination decision-making. Demographic characteristics were not systematically collected through a structured questionnaire. Some demographic information was available when participants disclosed this spontaneously during interviews. The first part of the interview focused on gaining insights into the vaccination decision-making process during adolescence. The second part of the interview focused on the decision-making process as an adult during the 18+ catch-up campaign. The conceptual model and interview guide (**Supplement 1**) were developed based on the Health Belief Model (HBM) and complemented using constructs from the Theory of Planned Behaviour and Betancourt’s model of Culture and Behaviour[18–20]. As such, the constructs this study explored were perceived susceptibility to and severity of HPV infection, perceived benefits and barriers of the HPV vaccine, cues to action, self-efficacy regarding HPV vaccination and sociocultural factors like cultural background, religious beliefs and social influence. The last part of the interview guide focused on current HPV vaccination implementation and ways to improve it. Before data collection, two pilot interviews were conducted to finetune the interview topic guide, validate the questions, and gain participant feedback. Data analysis was conducted concurrently with data collection to assess thematic saturation. Thematic saturation was reached after 16 interviews, as no further conceptual themes or relevant insights emerged by checking for new codes in the final three interviews. Written informed consent was obtained prior to the interview. The interviews lasted approximately 20-30 minutes and were recorded with the Philips LFH0648 voice tracer. The interviews were conducted in

Dutch. The audio recordings of the interviews were transcribed verbatim, without use of automated transcription software. Names and other identifiers were anonymised in transcripts.

### Data analysis

All transcripts were thematically coded using the software program MAXQDA (by PEK, LD, CJA, SvV). An initial coding framework was developed deductively based on the constructs of the conceptual model developed for this study[19, 20]. Two codes were created for each construct: reflecting vaccination decision-making during early adolescence and during the 18+ catch-up campaign. The transcripts were analysed using deductive thematic analysis based on this framework, with subcodes developed within the initial codes and additional codes identified inductively.

All interviews were coded by two researchers. Initial coding was conducted by PEK and LD, after which coded transcripts were independently reviewed by CJA and SvV. In addition, PEK reviewed and recoded interviews initially coded by LD to ensure consistency across coders. The coding framework was refined iteratively through discussions among the research team by modifying, merging, or removing codes to better reflect the data. All authors contributed to interpretation of the data and theme development. Coded extracts were compared to identify recurring themes. Coding was conducted in Dutch to preserve meaning and key excerpts were translated into English for reporting.

### Ethical considerations

The Medical Ethics Review Committee of Amsterdam University Medical Centers determined that the Medical Research Involving Human Subjects Act (WMO) does not apply, because participants were not subjected to actions or rules (reference number 2023.0343). Both the participant and researcher signed a consent form in duplicate prior to the interview.

Participants’ Audio recordings and transcripts were stored using pseudonymized participant identifiers. Transcripts and questionnaire data will be stored on secure GGD Amsterdam servers for a period of 10 years, in accordance with institutional data-retention policy. Only the research team will have access to the data.

## Results

### Participants’ characteristics

Sixteen women were included; twelve were recruited at HPV vaccination sites and four via snowball sampling. All experienced non-uptake of HPV vaccination during early adolescence and received HPV vaccination during the 18+ catch-up campaign. Participants were aged 19-27 years.

### Factors influencing HPV vaccination decisions during early adolescence and young adulthood

The identified themes that were mapped onto the conceptual model included sociocultural factors, perceived threat, evaluation of vaccination, self-efficacy and cues to action.

#### Social and cultural influences

Sociodemographic characteristics were not systematically collected. However, several participants shared characteristics that they considered relevant to HPV decision-making, including religious beliefs, migration background, and an affinity with alternative or natural-lifestyle beliefs. These themes are described below.

**During early adolescence**, non-uptake of HPV vaccination was strongly shaped by parental decision-making and broader parental sociocultural norms surrounding health, sexuality and vaccination. Twelve out of sixteen participants reported that their parents made the decision back then.

The association of HPV vaccination with sexual activity was reported as an important barrier for vaccination. Participants described how concerns regarding sexual activity reduced parents’ perceived relevance of HPV vaccination at a young age. These concerns were shaped by cultural taboos surrounding sexuality within community contexts (**Quote 1**), as well as religious beliefs regarding premarital sex (**Quote 2**).

> *1. “In Non-Western communities, it’s still somewhat taboo to talk about it [sexual activity] (…) My mother did receive that letter in the past, but she basically threw it away because she also thought, ‘Well, that’s not really necessary.’”*
>
> *2. “[Because of my religious beliefs], I was not planning to get the HPV vaccination. It’s more about not having sex before marriage and assuming I will only be with the person I marry.”*

Another participant described how her parents adhered to a holistic worldview, in which vaccines were considered unnatural and childhood illness was viewed as a part of normal development (**Quote 3**).

> *3. “Within [my parents’ philosophy of life], there is also very much the idea of vaccinations… things children go through are part of their development and vaccines are something unnatural.”*

Negative experiences of HPV vaccination, such as side effects experienced by peers reinforced hesitancy (**Quote 4**).

> *4. **“**I know that there were two girls who had received the vaccination and were in terrible pain in their arms, so it was really blown out of proportion, claiming that it was quite an intense vaccination.”*

**During the 18+ catch-up campaign**, sociocultural influences remained relevant but shifted in nature. Parental influence became less dominant (**Quote 5)**, while peer norms and broader social framing became increasingly important (**Quote 6**).

> *5. “When I asked around, everyone always seemed to have gotten it. Then I thought, ‘Gosh, why didn’t I…’”*
>
> *6. “Yes, I have talked about it [that I will get vaccinated] with them [my parents]. Their opinion has changed somewhat—they do have more knowledge about it now—but they’re still quite hesitant. They tend to lean more towards a ‘natural lifestyle’, so I thought, fine, so be it.”*

Stigma surrounding HPV vaccination appeared to decrease in young adulthood, as participants described the vaccine as less socially sensitive and less strongly associated with sexual activity than during early adolescence. This was partly attributed to the vaccine no longer being framed as a girls-only intervention, but increasingly as a general preventive health measure (**Quote 7**).

> *7. “It’s not only for girls anymore; when it was selective like that, it became more loaded, almost a taboo.”*

#### Perceived threat of HPV infection

**During early adolescence**, perceived susceptibility to and severity of HPV were generally low. Participants often indicated that neither they nor their parents perceived HPV as personally relevant, mainly due to their young age and presumed lack of sexual activity (**Quote 8**). Other participants reported low perceived susceptibility, as HPV was not a disease that was known in the family’s medical history (**Quote 9**).

> *8. “I wasn’t sexually active at all back then, so I really didn’t have any interest in it at all.”*
>
> *9. “Although there is a risk of cervical cancer, I still do not think my personal risk is high, as it doesn’t run in my family”*

**During the 18+ catch-up campaign**, most participants considered their susceptibility as high, often as they had initiated sexual activity or because friends tested positive for HPV (**Quote 10**).

> *10. “Several of my friends tested positive for HPV, so it sounds way more, how should I phrase it, closer to home that it could go wrong”*

Perceived severity also increased over time, driven by increased awareness and knowledge, fear of HPV-related disease (**Quote 11**), or personal experiences of a close relative being diagnosed with cervical cancer (**Quote 12**).

> *11. “The illness caused by HPV, the fear of cervical cancer, the stress surrounding it, and the concern that I might still develop it were decisive for me.”*
>
> *12. “I became much more aware after [a close relative] had cervical cancer [recently]. Knowing that HPV contributes to its development made me realise the seriousness of the virus and the benefits of vaccination. When I was younger, I received the invitation but didn’t really think about what HPV could cause. Experiencing it up close had a real impact on me, and that ultimately led me to get vaccinated.”*

#### Evaluation of benefits and barriers of HPV vaccination

**During early adolescence,** many participants recalled the novelty of the vaccine back then as a major barrier to vaccination, due to the lack of long-term safety and efficacy data at the time (**Quote 13**). Four participants reported that they had even been advised against vaccination by a medical doctor due to its novelty (**Quote 14**).

> *13. “It was the first time the vaccine was used in women; I personally found it very unsettling, because its consequences were not yet clear at that time.”*
>
> *14. “The general practitioner told me, ‘Well, just wait a little while’, and then this catch-up campaign started, so I thought, ‘Well, I’ll do it now’, because there was more research and more clarity about what the consequences were, and so on.”*

Fear of side effects was often based on anecdotal reports, media narratives or social networks and included chronic fatigue, infertility, paralysis and even death (**Quote 15**).

> *15. “And there was also an article published at the time, in which even a girl had died as a result of the vaccine.”*

These uncertainties and perceived risks outweighed the perceived benefits of vaccination during early adolescence. The only benefit one participant could recall was the belief that it could provide protection against HPV infection (**Quote 16**):

> *16. “In principle, if all went as it was supposed to do, it should protect against the virus, but that was about as far as my understanding went.”*

**During the 18+ catch-up campaign**, benefits clearly outweighed the barriers. Some already had the intention to get vaccinated before the campaign, but the costs were a barrier; the 18+ catch-up campaign then came as a good opportunity to get vaccinated for free (**Quote 17**).

> *17. “At one point I looked into it [getting the vaccination] and saw that it was quite expensive, so I thought, never mind. And then the campaign came and I thought, this is really a great opportunity to do it.”*

Trust in science and institutions further strengthened perceived benefits. Several participants indicated a generally positive attitude towards scientific developments and vaccination, which increased their willingness to accept the HPV vaccine. Others highlighted increased confidence due to the longer availability of the vaccine and accumulated research evidence (**Quote 18**).

> *18. “We’re now ten years further on, so much more research has been done… I have much more confidence that many more people have had the vaccine, so I don’t have any negative ideas about it anymore.”*

Experiences during the COVID-19 pandemic also contributed to this trust, illustrating the perceived effectiveness of vaccines more broadly (**Quote 19**):

> *19. “I think COVID also played quite a big role, because I really saw the impact a vaccine can have and how effective it is.”*

Finally, accessibility and convenience enhanced the perceived benefits of vaccination. Easily accessible vaccination locations, such as walk-in or pop-up sites, lowered the barriers and made uptake more attractive (**Quote 20**).

> *20. “I was just walking through the city and saw a sort of pop-up location where you could walk in and out without an appointment—that’s really accessible.”*

**During the 18+ catch-up campaign,** some barriers to vaccination remained. Some participants described psychological barriers, such as general hesitation towards vaccination (**Quote 21**)

> *21. **“**Well, maybe… it’s not really how I feel, but more that little voice saying: it is a vaccination, you don’t really know what it is.”*

Also, one participant questioned the personal relevance or added value of vaccination, for example in relation to uncertainty about already acquired HPV infection (**Quote 22**).

> *22. **“**I don’t know [whether I may already have been exposed to HPV], so whether vaccination will actually work for me.”*

#### Self-efficacy

**During early adolescence**, many participants questioned the capacity of young girls to make decisions on vaccine acceptance as they are minors. They reported low confidence in their ability to make an informed decision due to their age and limited knowledge (**Quote 23**) and given the complexity of the topic and its link to sexual health.

> *23. “I don’t know how actively you can participate in decision-making when you’re so young. Well, I might have… we did have real conversations about it. So, they did try. But you have no idea when you’re [that young].”*

**During the 18+ catch-up campaign**, self-efficacy among the participants increased. Participants attributed this to their increased age (**Quote 24**) and improved understanding of HPV and the vaccine.

> *24. “As a [young adolescent] you can’t really make that decision because you have no idea. As [a young adult] I can now make a very good decision about it.”*

#### Cues to action

**During early adolescence**, cues to action were primarily negative or passive, including rumours, negative media stories and discouraging advice from medical doctors, leading to non-uptake of vaccination. Although official invitation letters for vaccination were often reported as received, these did not function as an active trigger for vaccination uptake.

**During the 18+ catch-up campaign**, cues to action were more dynamic and reinforcing, creating a sense of urgency and accessibility. Although negative media stories and rumours were not necessarily absent, their influence appeared less salient, due to a greater number of positive and activating cues. Multiple simultaneous triggers, including formal reminders, media campaigns and social cues, such as friends also being vaccinated, reinforced decision-making (**Quote 25**).

> *25. “I think via TV ads, through advertising. And on Instagram all those things that keep popping up. And by talking about it with friends of course, like ‘are you getting it, have you already got it?’”*

The accessibility due to easy appointment systems, multiple locations to get vaccinated including locations that did not require an appointment, facilitated uptake (**Quote 20 and 26**).

> *26. “I saw the bus [vaccination bus to get vaccinated without appointment] and just thought I’m going to do it now.”*

### Recommendations for future HPV vaccination programmes

Based on these findings, several recommendations can be made for future HPV vaccination campaigns, which may increase uptake in the Netherlands.

First, communication strategies should provide clear, transparent and accessible information about HPV, vaccine safety and effectiveness (including specific information after sexual onset). Several participants indicated that communication should address common factors that are known causes of hesitancy directly, including concerns about side effects. Information should be in formats that are accessible and engaging for young people and parents. Several participants indicated that current governmental information was either too simplistic or too complex without presenting a holistic picture of both pros and cons (**Quote 27**).

> *27. “Either it was a very simple website without evidence-based information, or it was extremely complicated. There was not really anything in between. People want accessible but evidence-based information. And information that presents both sides as well, as there are also people who are really against injections.”*

Second, maintaining low-threshold and highly accessible vaccination services appears essential. Participants consistently described convenient vaccination service, including walk-in opportunities, pop-up locations and simple appointment systems as important facilitators for uptake during the 18+ catch-up campaign (**Quote 20, 26 and 28**).

> *28. “If you have to make an appointment somewhere far away in two weeks… no, you just want it now. I think especially this generation want things quickly.”*

Third, repeated reminders and visible media campaigns were perceived as effective triggers for action during the 18+ catch-up campaign. Several participants did not know they could get vaccinated for free up to the age of 18 (**Quote 29**). Participants therefore recommended repeated reminders during early adolescence, particularly around the age when young people become sexually active (**Quote 30).**

> *29. “As a child I did not know that I could still get the vaccine until age 18.”*
>
> *30. “I would definitely send invitations more often. Especially around the age when people become sexually active. So, I would target people around age 16, around the period of sexual education.”*

Finally, these findings strongly support the value of structural free-of-charge catch-up vaccination opportunities beyond age 18, as they may reconsider HPV vaccination later in life and can make independent healthcare decisions above 18 years old. (**Quote 31**).

> *31. “At a later age it would also be nice to still be able to get it for free. Actually, you should always have that right, even after age 18.”*

## Discussion

This study demonstrates that early-life non-uptake of HPV vaccination does not necessarily result in persistent non-uptake. The transition from non-uptake during early adolescence to acceptance in young adulthood was shaped by increased perceived susceptibility to HPV, greater autonomy in decision-making, improved confidence in vaccine safety and accessibility of the vaccination locations.

The finding that parents play a central role in vaccination decisions during early adolescence is well established in the HPV literature[12, 21, 22]. During early adolescence, participants reported limited self-efficacy and relied heavily on parental judgement, which was often shaped by cultural or religious beliefs, low perceived susceptibility and concerns about vaccine safety due to its novelty. Lower vaccine uptake among groups with affinity for anthroposophical or “natural” lifestyles has previously been described in the Netherlands[15, 23]. Our findings add nuance to this literature by showing that such early-life decisions do not necessarily result in persistent non-uptake. Rather, positions towards vaccination can change as individuals reassess earlier choices based on new social contexts, experiences and perceived risks.

The shift from parental influence during early adolescence toward peer influence in adulthood aligns with broader literature on health behaviour, suggesting that people increasingly orient themselves towards peer norms as their primary social reference in young adulthood[24]. Friends’ vaccination behaviour, peer conversations and experiences with HPV within social networks contributed to normalising vaccination and increasing perceived relevance. This shift reflects increasing autonomy and changing social reference groups during young adulthood, when individuals become more independent in healthcare decision-making. This transition has important policy implications. Since many participants experienced HPV vaccination non-uptake during early adolescence as a parental decision rather than their own, structural catch-up vaccination beyond age 18 may provide an important second opportunity for young adults to make an informed vaccination decision independently.

The association of HPV vaccination with sexual activity emerged as an additional barrier during early adolescence, particularly among participants who described religious beliefs or family and community norms in which discussions around sexuality were considered sensitive or taboo. In these contexts, HPV vaccination was sometimes perceived as less relevant at a young age or as indirectly linked to expectations around sexual behaviour. Similar concerns have been reported internationally, including in Middle Eastern, North African and Asian contexts, where HPV vaccination may be perceived as conflicting with norms regarding premarital sex or where girls are considered at low risk of HPV infection[25, 26]. Participants described the vaccine in young adulthood as a broader preventive health measure rather than something primarily associated with sexual behaviour. This shift in perception may contribute to reduced stigma and greater acceptability, although our findings do not allow conclusions about the effectiveness of specific communication strategies.

Perceived susceptibility to HPV infection emerged as a central driver of vaccine acceptance in adulthood, shaped by increased age, sexual debut and experience with HPV-related disease within the social network. These findings align with the HBM, which holds that health behaviour is influenced by perceived susceptibility and severity, alongside perceived benefits and barriers[19]. Previous HBM-based studies consistently showed that high perceived threat and high perceived benefits are associated with vaccine acceptance, for HPV[27–29] and for other vaccines, including COVID-19 vaccines[30]. While the HBM provided a useful framework for collecting and interpreting these findings, the model is less explicit in addressing how social relationships and life-stage transitions dynamically influence vaccination behaviour across the life course, all of which played a significant role in shaping vaccination decisions in this study. These results illustrate the dynamic nature of vaccine decision-making across the life course, shaped by shifts in social influences and increasing autonomy in young adulthood.

Barriers to vaccination during early adolescence, typically as perceived by the parents, included the lack of long-term safety and effectiveness information due to the novelty of the vaccine at the time of implementation. This uncertainty was not only shaped by public debate and media attention, but was further amplified by the fact that some clinicians and researchers in the Netherlands publicly expressed reservations about the national HPV vaccination policy at the time, which may have contributed to public doubt regarding vaccine safety. This is consistent with earlier Dutch research conducted during the introduction of the HPV vaccine in the Netherlands, when public debate and media attention contributed to uncertainty about vaccine safety [31, 32]. Over time, these concerns decreased, finally leading to vaccination acceptance among participants.

Participants reported increased trust in the HPV vaccine due to the accumulation of scientific evidence and its long-standing inclusion in the Dutch NIP. This reflects a broader pattern observed in vaccine acceptance[33, 34], whereby initial uncertainty surrounding new vaccines decreases as more data and real-world experience become available.

These findings should be interpreted within the Dutch context, where HPV vaccination has been characterised by moderate uptake and context-specific forms of hesitancy. Early public debate during the introduction of the HPV vaccine in the Netherlands, together with the presence of religious and anthroposophical communities, has shaped the salience of concerns about vaccine safety, natural lifestyle preferences and moral implications. These concerns may vary in prominence across European contexts. These findings also reflect the broader context in which HPV vaccination decisions are made, with some participants describing culturally mediated concerns during childhood decision-making. The changing nature of these concerns in adulthood suggests that such beliefs may be less stable than is often assumed and that their influence may depend on life stage, social environment and the availability of alternative frames, such as cancer prevention and personal health responsibility.

This study has several limitations. Participants were asked to retrospectively reconstruct their childhood decision-making, which could introduce recall bias. Memories of reasons for non-uptake a decade earlier may have been reshaped by subsequent experiences and current attitudes. In addition, given the strong influence of parents at the time, some participants may have assumed rather than directly recalled their parents’ reasoning. Participants’ accounts of parental reasoning are necessarily mediated through childhood memory and may be influenced by current rationalisations.

The study was conducted in Amsterdam, a highly urban and multicultural setting, which may limit generalisability to other regions of the Netherlands where cultural and religious dynamics differ. Detailed demographic characteristics were not systematically collected using a structured questionnaire. Although several participants spontaneously shared relevant information regarding cultural background, religious beliefs, or family context during interviews, these data were incomplete and not available for all participants. This limited our ability to assess how demographic characteristics such as migration background, education, socioeconomic status, or religion systematically shaped HPV vaccination decision-making across participants. Finally, and most importantly, only women who ultimately chose to receive the HPV vaccine were included. As such, the analysis does not provide insights into factors influencing individuals who continue to decline vaccination.

## Conclusion

This study illustrates that HPV vaccine acceptance is not a static state. It is dynamic and responsive to shifting personal, social and structural circumstances over time. The life-course changes documented here (autonomy, perceived risk, peer influence) are predictable developmental transitions, making a standing permanent catch-up offer a rational policy tool. Within the Dutch setting, such a catch-up could be achieved, for example, by offering vaccination at an older age in settings like the Centres for Sexual Health, or at a younger age within existing Youth Health Care (by sending active reminders while HPV vaccination is still free before the age of 18).

## Declaration of Interest Statement

The institution of MFSvdL, CJA and SvV received study funding for an investigator-initiated study from GSK; MFSvdL served on advisory boards of MSD and Novosanis; payments were made to his institution. This disclosure is made for transparency purposes and does not imply a conflict of interest with the present publication, which is unrelated to the grant’s scope and does not receive support from it. Other authors declare that they have no conflict of interest.

## Author Contributions

CJA, MFSvdL and JAB contributed to the conceptualization of the study. LD, CJA, MFSvdL and JAB were involved in the methodology and preparation of the non-WMO approval. LD and PEK were responsible for data collection (interviews). LD, PEK, CJA, SvV and EW contributed to data curation and formal analysis. AW supported investigation and facilitated data collection. UD and GNN contributed to the interpretation of data and provided input on the analytical approach. SvV, PEK and CJA were responsible for writing the original draft. All authors contributed to the review and editing of the manuscript and approved the final version.

## Data Availability

Data Availability Statement
All data produced in the present study are available upon reasonable request to the authors.

## Acknowledgements

We would like to thank all participants for sharing their experiences and perspectives in this study. We also thank the staff involved in the organisation of the HPV vaccination campaign and participant recruitment in Amsterdam. Dr C.J. Alberts acknowledges support from her personal VENI fellowship, ZonMw, the Netherlands (grant number 09150162210124).

## Data Availability Statement

All data produced in the present study are available upon reasonable request to the authors.

## Notes

### Author Declarations

The Medical Ethics Review Committee of Amsterdam University Medical Centers determined that the Medical Research Involving Human Subjects Act (WMO) does not apply, because participants were not subjected to actions or rules (reference number 2023.0343).

