## Supplement 1 for "From non-uptake during early adolescence to acceptance during a catch-up campaign: a qualitative study of changing HPV vaccination decisions across two life stages among young women in Amsterdam, the Netherlands"

a) Department of Infectious Diseases, Public Health Service of Amsterdam (GGD Amsterdam), the Netherlands; b) Department of Epidemiology and Data Science, Amsterdam University Medical Center, Amsterdam, The Netherlands; c) Amsterdam Institute for Immunology and Infectious Diseases (AI&I), Amsterdam UMC, University of Amsterdam, Amsterdam, The Netherlands; d) School of Nursing, Midwifery, Allied and Public Health, Canterbury Christ Church University, North Holmes Road, Canterbury, Kent, CT1 1QU, United Kingdom; e) Department of Healthy Living, Public Health Service of Amsterdam (GGD Amsterdam), the Netherlands; f) Department of Social Psychology, University of Amsterdam, Amsterdam, the Netherlands; g) Amsterdam UMC location University of Amsterdam, Department of Internal Medicine, Amsterdam, The Netherlands; h) Amsterdam Public Health Research Institute (APH), Amsterdam, the Netherlands; i) Cancer biology and immunology (CCA) Research institute

**Supplement 1: Interview guide**

**Part 1: Decision Back Then (5-6 minutes)**

1. Can you tell me how the decision was made not to get the HPV vaccination when you were invited at the age of 12?

*Possible follow-up questions. Depends on the factors mentioned in question 1*

| | |
|---|---|
| **Perceived susceptibility***<br>Can you tell me what you knew about the HPV virus when you were 12 years old?<br>How did you think HPV was spread?<br>Who did you think could get HPV?<br>Did you think at that time that you could get it? | **Perceived severity***<br>Can you tell me what you knew at that time about the consequences of getting an HPV infection?<br>Did you know what the consequences are of having an HPV infection?<br>How serious did you think an HPV infection is/can be?<br>In how many cases did you think it could have severe consequences? |
| **Perceived benefits****<br>Can you tell me what you knew at that time about the HPV vaccination?<br>Did you know the vaccination existed?<br>Did you know you could get it (for free)?<br>Did the vaccine seem useful to you?<br>What were the reasons at that time that might have influenced you to consider getting the vaccination?<br>How did these reasons play a role in your decision not to take the vaccination?<br>Which factors weighed more heavily, which less? Why? | **Perceived barriers****<br>What were the possible disadvantages or barriers for you at that time to get the vaccination?<br>Were there risks?<br>Were there logistical problems?<br>How did these disadvantages or barriers play a role in your decision not to get vaccinated at that time?<br>Which factors weighed heavily, which less? Why? |
| **Self-efficacy**<br>To what extent did you feel capable at that time of deciding about getting the vaccination?<br>Where did you find your information?<br>Did you find that information useful enough? | **Cues to action**<br>Were there external factors at that time that prompted you to consider getting the vaccination? Think of the RVP invitation, campaigns, or a healthcare worker who talked about it |

| | |
|---|---|
| Did you miss information? What kind of information did you miss? How would this information have helped you make a different decision? | How did these factors contribute to making a decision? |
| **Social norms**<br>What was the opinion of people around you about the HPV vaccination?<br><br>Did the opinions of other people play a role in your decision at that time? If so, in what way?<br><br>Did your parents play a role in this decision? If yes, how was their influence divided among them? Did one have a greater role than the other? How and why? What was the role of the other? | **Cultural factors**<br>Are there other (cultural) factors that influenced your decision at that time? (Think about culture or religion) |

**perceived susceptibility + severity = perceived threat; ** benefits + barriers = evaluation of behavior*

2. To what extent did you see HPV as a threat to you? Did this influence (how) you made the choice at the time?
3. What were the main pros and cons of vaccination at that time that led to the decision not to get vaccinated?
4. Which factors we just discussed were the most important in making that decision?
   *End part 1*

**Part 2: the choice now (5-7 minutes)**

5. Can you tell me how the decision was made now to vaccinate against HPV?

| | |
|---|---|
| **Perceived susceptibility***<br>Can you tell me what you currently know about the HPV virus?<br><br>How do you think HPV is spread?<br><br>Who do you think can get HPV?<br><br>Do you think you are at risk of getting HPV? | **Perceived severity***<br>Can you tell me what you know about having an HPV infection and its possible consequences?<br><br>Do you know what the consequences are of having an HPV infection?<br><br>How serious do you think an HPV infection is/can be?<br><br>In how many cases do you think it is severe? |
| **Perceived benefits****<br>Can you tell me what you now know about the HPV vaccination? | **Perceived barriers****<br>Do you see any disadvantages or barriers to getting the HPV vaccine? |

| | |
|---|---|
| What do you think the vaccine protects against?<br><br>How effective do you think the vaccine is?<br><br>What are the reasons/benefits for you now to have taken the vaccine?<br><br>How did these benefits play a role in your decision to take the vaccination now?<br><br>Which factors weigh heavily in this decision, and which weigh less? | Are there risks?<br><br>Are there logistical problems?<br><br>Which disadvantages played a role in your decision?<br><br>Which factors weighed heavily in this decision, and which weighed less? |
| **Self-efficacy**<br>To what extent do you feel capable of making an informed decision now?<br><br>Where did you find your information?<br><br>Did you find that information useful enough?<br><br>Did you miss any information? | **Cues to action**<br>Are there other external factors that prompted you to take the vaccination?<br><br>Think about the RVP invitation<br><br>Think about campaigns<br><br>Think about a healthcare worker who talked about it<br><br>How did these factors contribute to making a decision? |
| **Social norms**<br>What is the opinion of people around you about the HPV vaccination? Think about friends, family, etc.<br>Did other people's opinions play a role in your decision to get the vaccination? If so, how? | **Cultural factors**<br>Are there other (cultural) factors that were important in making the decision now? (Think about culture or religion) |

**Perceived susceptibility + severity = perceived threat; ** benefits + barriers = evaluation of behavior*

6. To what extent do you perceive HPV as a personal threat? Did this influence your decision now?
   - If high: Why do you consider HPV a threat? What are you afraid of? (HPV or cancer)
   - How does this compare to before being vaccinated?
7. What are the most important pros or cons of the vaccine for you now?
8. Which factors we just discussed were most significant in making that decision?

*End part 2*

**Part 3: the difference between then and now (5-7 minutes)**

Name the factors she mentioned earlier and compare them with what she says now.

10. What do you consider the most significant difference between then and now in making the decision?
11. Why/How has this changed compared to before?
12. Would you have wanted to be more actively involved in the decision-making process? How? Why?
13. How do you look back on the process now and receiving the vaccine at a later age?

*End part 3*

**Part 4: the policy (~5 minutes)**

14. Why do you think the HPV vaccination rate is lower than that of other vaccines?
    - o Explain what the policy is now. What do you think of this policy?
    - o What do you like about this policy?
    - o What do you think could be improved about this policy? How?
    - o Do you think the government should do more to increase the HPV vaccination rate?

15. What is your opinion on the catch-up campaign in which you received the HPV vaccination now?
    - o What went well during this catch-up campaign?
        - o What do you think could be improved about this campaign? How?*End part 4*

END: These were the interview questions. We have reached the end of the interview. Is there anything you would like to add? Anything important that hasn't been discussed but you consider significant? Do you have any further questions? Any questions about the topic or the interview?

Thank you for your help and time!
